# Potentials of constrained sliding mode control as an intervention guide to manage COVID19 spread

**DOI:** 10.1101/2020.09.21.20166934

**Authors:** Sebastián Nuñez, Fernando A. Inthamoussou, Fernando Valenciaga, Hernán De Battista, Fabricio Garelli

## Abstract

This work evaluates the potential of using sliding mode reference conditioning (SMRC) techniques as a guide for non-pharmaceutical interventions and population confinement to control the COVID-19 pandemic. SMRC technique allows robustly delimiting a given variable in dynamical systems. In particular, for the epidemiological problem addressed here, it can be used to compute day by day the contact rate reduction requirement in order to limit the intense care units occupancy to a given threshold. What is more, it could impose a given approaching rate to the health care system limits. Simulations are performed using the well-known SEIR model fitted to the Argentinian case to demonstrate what this control strategy suggests, while the effect of realistic period transitions between different confinement levels are also considered.

## 1 Introduction

Since the end of 2019, a novel coronavirus (SARS-CoV-2) started spreading around the world. The disease quickly turned into a worldwide health crisis, leading to the World Health Organization to declare the COVID-19 infection a pandemic on March 11th, 2020. Until the submission of this work the COVID-19 disease is affecting 213 countries and territories with more than 25 million reported cases and more than 800 thousand deaths [1].

Vaccines and effective treatments are under development. In the meanwhile, the main strategy to deal with the COVID-19 outbreak has been the implementation of non-pharmaceutical interventions (NPIs). Among them, people confinement and social distancing have been imposed to attenuate the number of infected individuals (i.e. to “flat the curve”), with the aim of avoiding the saturation of the health systems. In particular, intense care unit (ICU) occupancy levels represent the bounds the governments are more worried about. The NPIs strategies include school and universities closures, social event bans, border closures, work bans for non-essential activities, social distancing, quarantines and lockdowns [2]. Isolation of confirmed cases including tracing and testing the close contacts contributes in reducing the community transmission of the virus. Additionally, the use of technology could provide early identification and monitoring of cases [3].

In Argentina, the first case was reported on March 3rd, 2020. From March 12th, travelers arriving from outside the country were sent to mandatory quarantine. Then, on March 15th a stay-at-home recommendation was emitted including closing schools and universities. Borders were closed on march 16th and other activities like sport and social events were banned. On march 19th, a lockdown was established with exceptions given for workers related to essential activities. This lockdown was extended during the first weeks of April. From then on, the order was lift in those regions of the country less affected and some relaxations were implemented in Ciudad Autónoma de Buenos Aires and its surrounding area (within the region known as AMBA). As of September 1st, more than 400 thousand cases have been reported, including more than 8 thousand deaths [1]. The last available data show that a high percentage of new infections is concentrated in the AMBA. However, as the economical activities are resumed and mobility constraints are relaxed, the number of reported cases in some provinces is increasing.

Epidemiological models have been applied to the analysis and forecast of the COVID-19 disease in many countries (see for instance [4, 5, 6]). The models also play a significant role in the evaluation of potential interventions and the problem of determining a suitable NPI policy has received much attention in recent months. Particularly, a variety of approaches have been proposed to the problem of designing NPI policies subject to constraints by applying tools from control theory. An optimal control problem for NPI design is formulated in [7] with the aim of reducing infected population and contaminated objects. In [8], optimal intervention for reducing the peak of infections is proposed. Contact tracing policy and hospitalization of infected cases are considered for application of the optimal control approach in [9]. In [10], age structure is considered for the design of quarantine rates of individuals. The application of a proportional controller is proposed in [11] in which the objective is to maintain the number of hospitalized individuals below a set-point. In [12], a dynamic optimization approach is utilized with the aim of minimizing a socioeconomic cost function subject to limiting peak value of infections. Model predictive control (MPC) was considered in [13] with the aim of designing an on-off NPI strategy. In [14] interval arithmetic is applied for the design of a robust MPC based feed-back that weekly updates the NPI policy. In [15] a cyclic exit strategy is proposed where a number of continuous days of work is followed by a number of days of lockdown. In [16] model-based intervention policies are determined with the aim of maintaining the system evolution constrained within a safe set. Then, predictors are utilized for estimating the states required in the NPI. The application of a bang-bang controller type is considered in [17], where the objective is to avoid the number of ICU beds is exceeded above a threshold value. Under certain conditions on parameters, it is shown that the closed-loop system has a global and bounded solution satisfying the constraint with finite jumps. In [18] a criterion for optimal NPI design with minimal duration taking into account health care system capacity is presented. Sliding mode (SM) control has also been considered for the design of vaccination strategies [19] and regulation of affected individuals [20, 21]. Particularly, for COVID-19 disease, a sliding-mode regime is proposed in [22] to limit the number of exposed individuals.

In this work, we periodically compute the required action of the NPI strategy to reach a given infectious threshold with a desired approaching rate. This computation is performed via a sliding mode reference conditioning (SMRC) scheme based on measurement of the infectious cases. Even though we consider a susceptible-exposed-infectious-removed (SEIR) epidemiological model to represent the spread of COVID-19 along the population, any other model could have been considered. Indeed, differing from other control approaches, the SMRC scheme is insensitive to model parameters variation provided a sufficiently fast (with respect to the system dynamics) measurement and actuation frequency is implemented (i.e. daily). A distinctive feature of the proposed strategy is that it is directly tuned by the two expected outcomes: the threshold on the infectious population and its corresponding approaching rate. From the setting of these intuitive parameters, the SMRC scheme automatically shapes the NPI action requirement to fulfill the desired constraints on the epidemic course. Given the particular case addressed here, issues concerning real-life implementation are also discussed and evaluated.

The rest of the paper is organized as follows. Section 2 describes the SEIR model and the control problem is stated. Section 3 presents the SMRC scheme for NPI computation. Section 4 shows numerical results under different scenarios. Section 5 discusses implementation issues and more realistic simulations are presented. In Section 6 conclusions and future work are outlined.

## 2 Model of infectious disease spread and problem statement

### 2.1 The SEIR epidemiological model

The approach utilized to represent the infectious disease dynamics is a compartmental model with four compartments: Susceptible, Exposed, Infectious and Removed (Fig. 1). The parameter *β* is the average number of contacts between individuals in the Susceptible compartment (S) with infectives per unit of time [23]. Then, (*βI/N*)*S* represents the number of new cases per unit time due to the S susceptibles which are removed from S and incorporated to compartment E. The individuals in E had contact with the disease but they are not yet infectious. After an average time of 1/λ days an individual in E is moved to the Infectious compartment. Finally, after an average number of 1/γ days it is moved to the Removed compartment where the case is no longer active (i.e. the individual is recovered or dead). It is assumed that an intervention policy can be implemented in a way that the contact rate between susceptible and infectious individuals can be diminished. Therefore, *β* can be replaced with (1 − *u*)*β* [16, 18], with *u* ∊ *[U_min_, U_max_*] and 0 ≤ *U_min_ ≤ U_max_ ≤* 1. This formulation leads to the following set of ordinary differential equations:

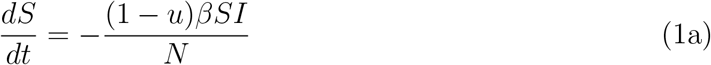

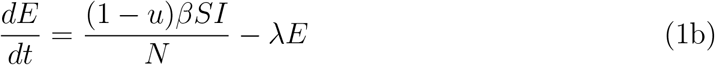

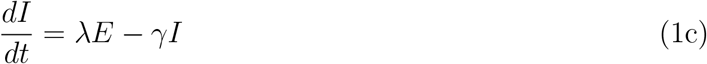

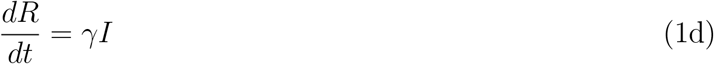

with non-negative initial conditions. Since a constant population was considered, eqs. (1) satisfy *S*(*t*) + *E*(*t*) + *I*(*t*) + *R*(*t*) = *N*. Additionally, the model can be normalized with respect to the size of the population (i.e. *s*(*t*) = *S/N*, *e*(*t*) = *E/N*, *i*(*t*) = *I/N*, *r*(*t*) = *R/N* represent the fraction of the population in each compartment), leading to a normalized version of model (1).

**Figure 1:**
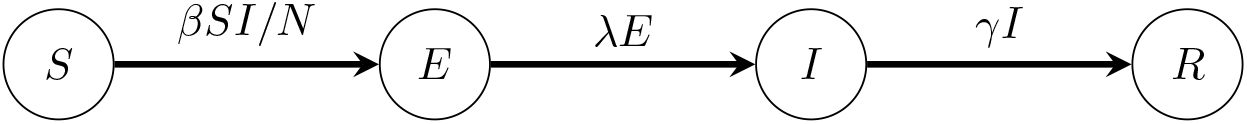
Schematic representation of the Susceptible, Exposed, Infectious and Removed (SEIR) compartmental model.

### 2.2 Problem Statement

If the number of infectious cases rises above certain critical level, the health care systems capacity may saturate. Then, as the number of available ICU is surpassed the quality of health provided to individuals deteriorates and consequently, the death counts may increase dramatically. In many countries, a variety of measures were taken with the aim of flattening the infection curve and ‘to slowing down’ the progression of the disease. Then, crucial time could be gained not only to incorporate new medical equipment but also to provide staff training. Based on this observation the following constraint can be formulated

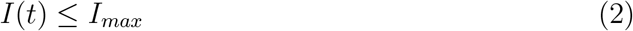

where *I_max_* is a value provided by authorities such that it ensures the health care system response will be adequate. Then, the design of *u*(*t*) as an NPI policy is required. Recalling that *u*(*t*) ∊ *[U_min_ U_max_*], a time varying function could be applied according to the disease time evolution.

## 3 Proposed control strategy

### 3.1 Sliding Mode Reference Conditioning algorithm

The proposed control scheme is based on the adjustment of the control input u according to the evolution of infectious individuals I. The idea is to fulfill constraint (2) and to reduce the risk of a collapse of the health care systems. To this end consider a continuous measurement of the infectious individuals. Then, the following auxiliary function is proposed

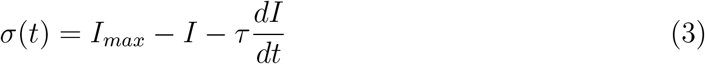

where *τ* > 0 is a design parameter. The proposed control action is:

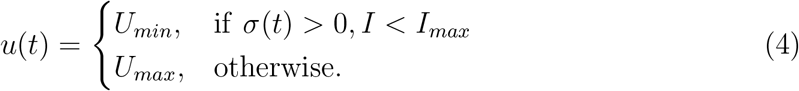

Assume the system starts at *I*_0_ *< I_max_*, since *σ* > 0, *u* = *U_min_* is applied. As the number of active cases increases the function in eq. (3) decreases and eventually it may try to cross below zero. If this is the case, the control action in (4) applies *U_max_* in order to modify the trajectory of *I*(*t*). Once *σ* is above zero, the control action applies *U_min_* again. In the ideal case, an SM regime is established and the system trajectory slides on *σ* = 0 thus preventing the number of infectious individuals above the selected level. It is important to remark that, differently from conventional sliding mode control, the sliding regime is not sought here. Instead, it is only established as a transient mode so as to avoid I surpassing the rate of change imposed by τ or the limit value *I_max_*. The sliding regime immediately finishes when the evolution of the infectious people does not exceed the imposed limits any more.

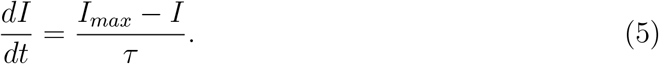

As the constraint *σ* = 0 is enforced to the system, the dynamics of I results in:

Thus, the parameter τ adjusts the approaching rate of *I* to *I_max_* and can be used to control the speed at which the rate of infectious cases tends to the limit. Fig. 2.A presents an example of the time evolution of *I* for different values of *I_max_*. The corresponding phase-plane plot is shown in Fig. 2.B, where the constraint *σ* = 0 corresponds to straight lines joining the points (0, *I_max_*/*τ*) and (*I_max_*,0). When the system trajectory reaches *σ* = 0, it slides on this constraint towards the point (*I_max_*,0) fulfilling eq. (2).

**Figure 2:**
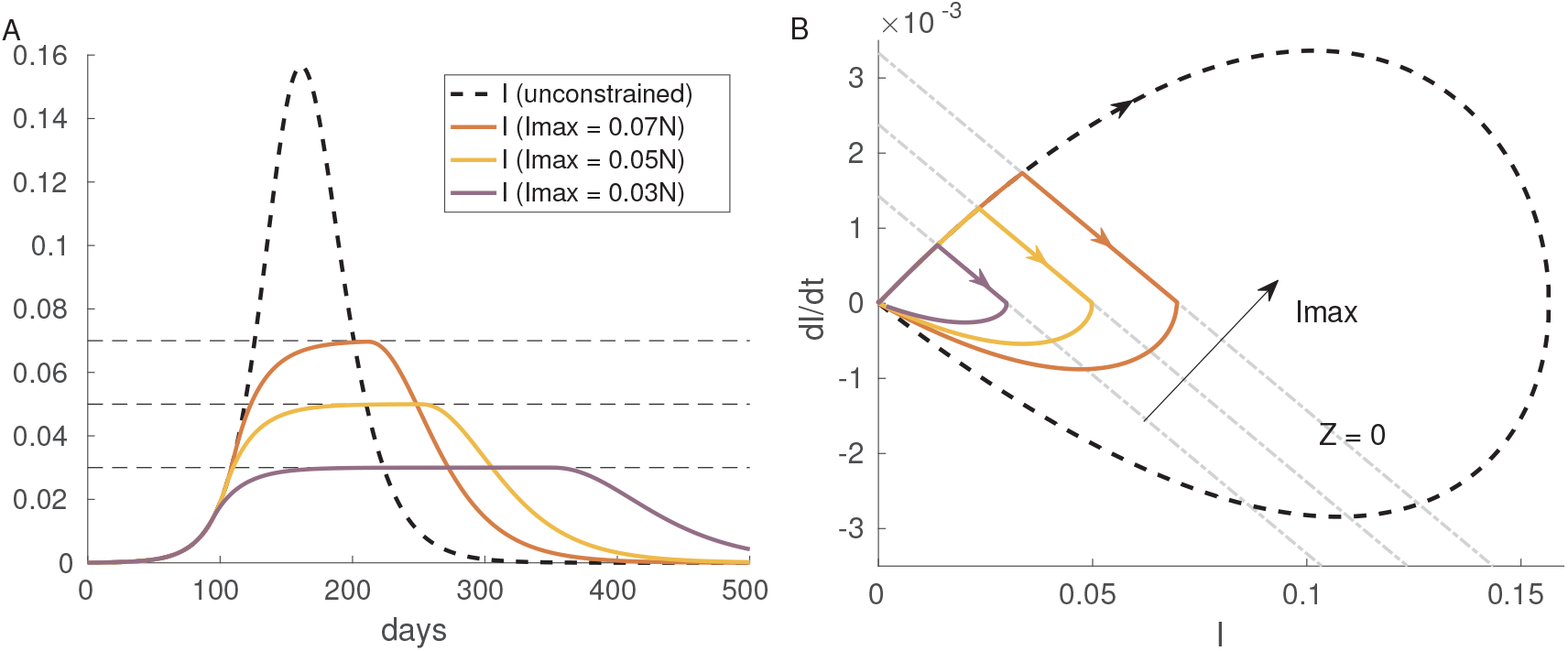
Illustrative simulation of the proposed controller with *τ* = 21 days. (A) time response of I at different levels of *I_max_* (B) phase-plane (*I,dI/dt*). Parameters: *β* = 0.17 day^−1^, *λ* = 1/5.1 day^−1^, *γ* = 1/14 day^−1^

### 3.2 SM existence and robustness conditions

A SM regime can be established only if there is a unitary relative degree of *σ*(*t*) with respect to the manipulated variable (the transversality condition [24]). By replacing *dI* /*dt* in eq. (3):

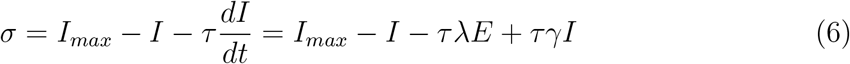

The time derivative of *σ* results in

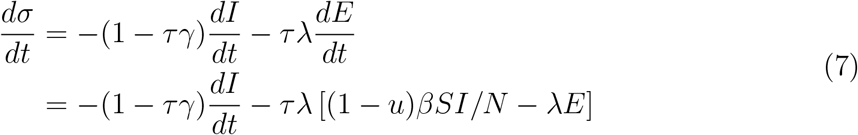

According to eq. (7) the relative degree of σ with respect to the control action u is unitary as long as the rate (*βSI*) is not zero.

Additionally, the control action must be high enough to enforce a sign change on the time derivative *dσ*/*dt* at *σ* = 0. The so-called equivalent control (*u_eq_*), a continuous equivalent signal that would lead to the same dynamics resulting from the SM (but without any of its robustness features), can be obtained by setting eq. (7) to zero:

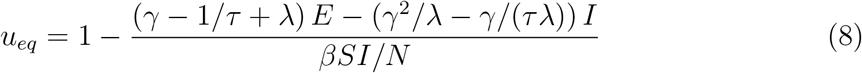

This expression must take values in the range [*U_min_, U_max_*] for SM to exist on *σ* = 0. Along this surface the number of exposed individuals is

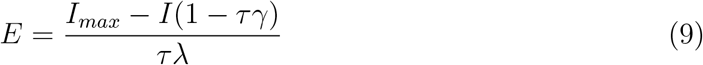

By replacing (9) in (8), it can be shown that the numerator of the second term is

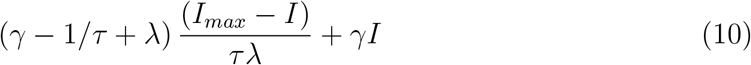

This term is positive in the region of operation of the SMRC (*I* < *I_max_*) and consequently *u_eq_* < 1. If *U_max_* < 1 is provided, eqs. (8)-(10) can be used to determine whether the SM regime exists. As *I*(*t*) → *I_max_* the required control action tends to 1 − 1/ (*βS*/*N*/*γ*), a singular control action that maintains *I*(*t*) = *I_max_* [18].

It is worth noting that *γ*, *λ* and the contact rate *β* in (7) are co-linear with the control action. That is, these important model parameters fulfill the matching condition [24]. Thus, provided the equivalent control action in (8) remains within the range [*U_min_*, *U_max_*] (i.e. the necessary and sufficient condition for SM holds), the scheme is insensitive with respect to variations on either *β*, *γ* or *λ*.

## 4 Results and discussion

First, and to show the proposal features, an ideal theoretical condition is assumed. Under this framework, the number of infectious is continuously reported and the ideal control law is applied, i.e., when *σ* = 0 is reached the control input can be switched according to eq. (4) at infinite frequency. Then, in Section 5 simulations for a more realistic scenario are compared with the ideal one to show the proposal applicability.

The parameters values utilized with the SEIR model were: *λ* = 1/5.1 day^−1^ and *γ* = 1/14 day^−1^. These values are in line with typical values for the disease [5]. Other parameters values were chosen as *N* = 2890151 and initial conditions (*I*_0_, *E*_0_, *R*_0_) = (10, 300, 0) and *S*_0_ = *N* − *E*_0_ − *I*_0_ − *R*_0_. Further, comparing the infectious level with the occupancy of ICU beds reported by the CABA government on 08/18/2020, with the total number of ICU beds being 450, *I_max_* = 1.54e5 limit was established. The assumption of a constant ratio within ICU patients and active cases is unrealistic as this ratio is decreasing. An improvement would be the incorporation of an ICU requirement estimator. Anyway, *I_max_* could be updated periodically according to availability in the health care system and ICU occupancy ratio variations.

### 4.1 Scenario 1

Two scenarios were simulated. The first one, in Fig. 3, assumes a *β*=0.22 day^−1^ which approximately corresponds to a disease evolution without any restriction or prevention policy (use of masks, reduction in people mobility, etc.). The subplots at the left column show, in a downward direction: the Susceptible (S) level, the Infectious (I) level, and the restriction policy level (1-u). The figure is completed with the Exposed (E) level and the phase-plane evolution (*I,dI*/*dt*) at the top and bottom of the right column, respectively. It is worth mentioning that the values that represent the sets S, E and I are plotted relative to the considered population N, and that a level of 1-u equal to 1 corresponds to no restriction at all, meanwhile a value of 0.6 corresponds to a restriction level of 40%.

**Figure 3:**
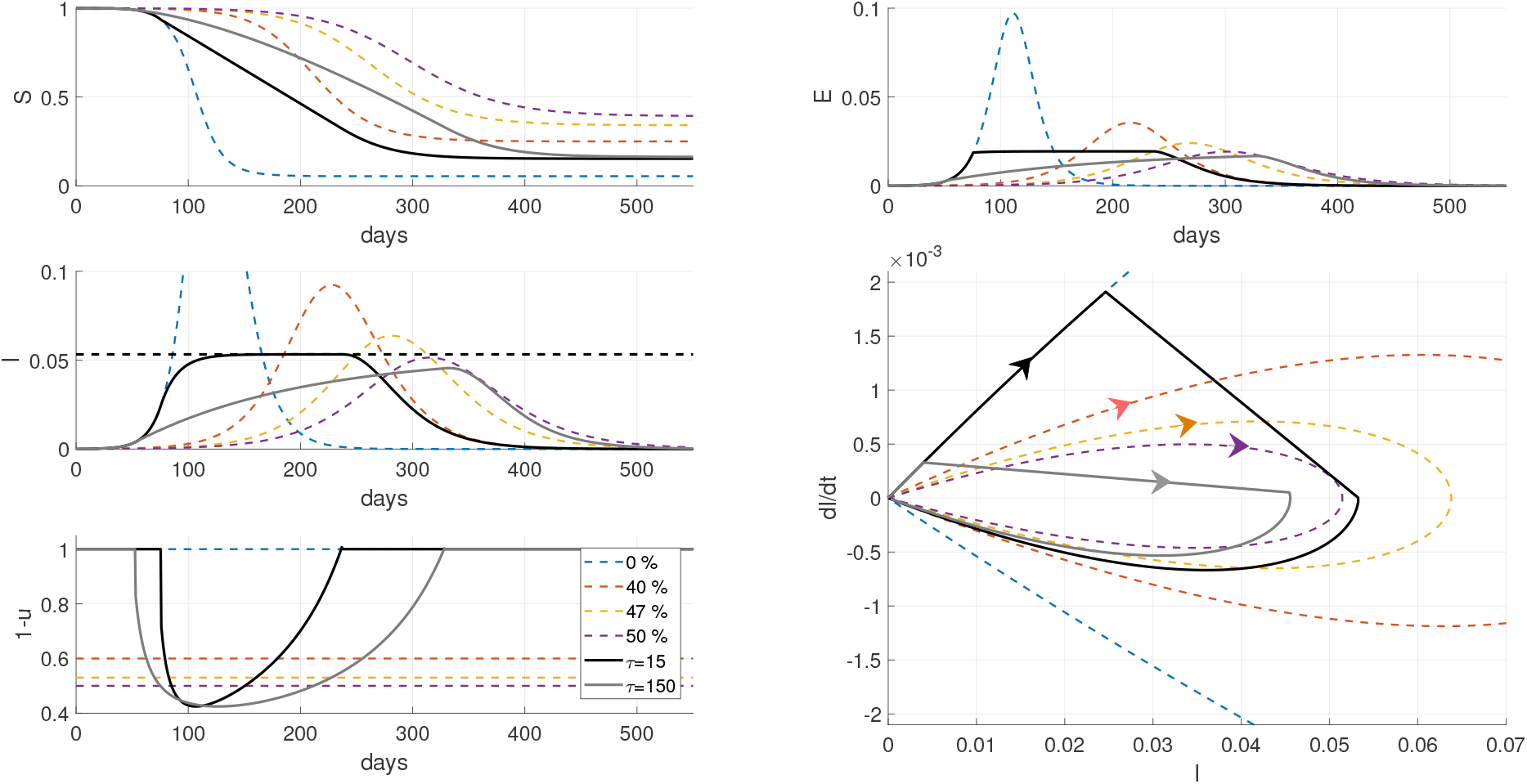
Scenario 1: *β*=0.22 day^−1^. Simulations with the SEIR model in a theoretical scenario where the switching control action (eq. (4)) is implemented. From left to right and top to bottom: time evolution of Susceptible (S), Exposed (E), Infectious (I), phasespace plot (I,dI/dt) and restriction actions (1-u). Colored dashed lines represent constant levels of restrictions. Solid black and gray lines depict the closed-loop evolution for *τ*=15

The disease evolution without any restriction level is depicted with the blue dashed lines in Fig. 3. In the same way, with orange, yellow and violet colors it is represented the system behavior for different levels of constant restriction, i.e, 40%, 47% and 50% respectively. As expected, the infectious peak decreases while the total epidemic duration increases as the restriction becomes harder. The solid lines in black and gray represent the SMRC closed-loop system evolution for *τ*=15 and *τ*=150 days, respectively. As can be seen, when the infectious level complies with the imposed limitation (both absolute value and approach speed), the restriction level begins to increase to avoid exceeding the imposed limit (horizontal dashed line in the infectious subplot). When this condition ceases to be fulfilled (due to the evolution of the disease) the restriction level begins to decrease until the level without restriction is recovered again. The main difference between the proposal behavior and the constant restriction levels is the non-saturation of the health system, which is only achieved for a constant restriction level above 50 %. Not exceeding the health system capacity brings with it the most important result of not increasing the mortality rate.

As can be appreciated from Fig. 3, the evolution of the closed-loop system for the case with *τ* =15 days has a similar duration to the case of 40% constant restriction (dashed orange lines). The main difference is the “time distribution” of infectious agents that avoids surpassing the health system capacity. This is achieved by keeping the system unrestricted for a longer time period, but requiring a deeper restriction peak. To make a comparison, the 40 % constant restriction case has an approximate duration of 400 days, while for the closed-loop system there exist restrictions during only 161 days. 101 of these days correspond to a restriction level lower than 40 %, whilst the remaining days the restriction is harder reaching a peak of 58 % for a few days. Then, the closed-loop evolution would allow obtaining 239 extra days without any type of restriction, which is an improvement of 60 %. With the idea of quantifying this feature, a performance index (Restriction Level Index, RLI) is proposed as:

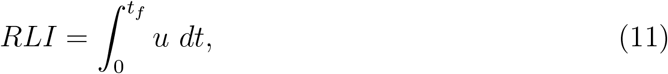

where *t_f_* corresponds to the total time duration of the epidemic. A higher value of RLI implies a harder restriction. As can be seen from Table 1, both closed-loop cases achieve a lower level of restriction and, as expected, the case with *τ* =150 increases this value over the *τ* =15 one.

**Table 1:** Performance index on applied restrictions

| Scenario 1 |  | Scenario 2 |  | Comparison |  |
| --- | --- | --- | --- | --- | --- |
| Case | RLI | Case | RLI | Case | RLI |
| 40% | 220 | 10% | 60 | 40% | 220 |
| 47% | 258 | 20% | 120 | 47% | 258 |
| 50% | 275 | $\tau=15$ | 4 | $\tau=15$ | 66 |
| $\tau=15$ | 66 | $\tau=150$ | 20 | $\tau=150$ | 263 |
| $\tau=150$ | 121 | - | - | - | - |

The solid gray lines in Fig. 3 show the system evolution for *τ* =150 days. This case would serve as a recommendation to be applied from the detection of the first confirmed case. As can be seen, the system behavior resembles the 50 % constant restriction case as far as disease duration is concerned. The use of a larger τ allows starting acting much earlier in the face of a rapid approach to the imposed limitation. As the figure depicts, the imposed limit is never reached obtaining a lower infectious peak than in the 50% constant restriction case, also with a shorter duration in restrictions.

The lower right box in Fig. 3 plots the phase-plane system evolution for all cases. As shown, the closed-loop ones first follow system evolution without restriction until the limiting condition is met. Then, the system evolves with the imposed dynamics (5), reflected in the diagonal straight-lines, towards the defined Imax level so as not to exceed the health system capacity. The SMRC adaptation becomes inactive once the risk of exceeding *I_max_* ceases.

### 4.2 Scenario 2

The second scenario, Fig. 4, assumes *β* =0.117 day^−1^ which approximately corresponds to the disease evolution under the restrictions and prevention measures implemented by the Argentinian government [25]: use of masks, restriction of mobility only to essential personnel, etc. The boxes distribution in the figure is the same as in the previous scenario. The system evolution for these parameters and without extra restrictions corresponds to the dotted lines in blue. The orange and yellow dotted lines represent the system behavior for a 10% and 20% extra constant restriction level respectively. Now, these levels of restriction must be understood as an additional level over the measures already applied, which are summarized in a *β*=0.117 day^−1^. The solid red line shows the disease evolution for real data published by the city government in [26] until 08/19/2020. As can be seen, the adjustment of the real data matches the system evolution for *β* =0.117 day^−1^ without any additional restriction. As shown, if the evolution of the disease followed this trend, the system would be saturated on day 248.

**Figure 4:**
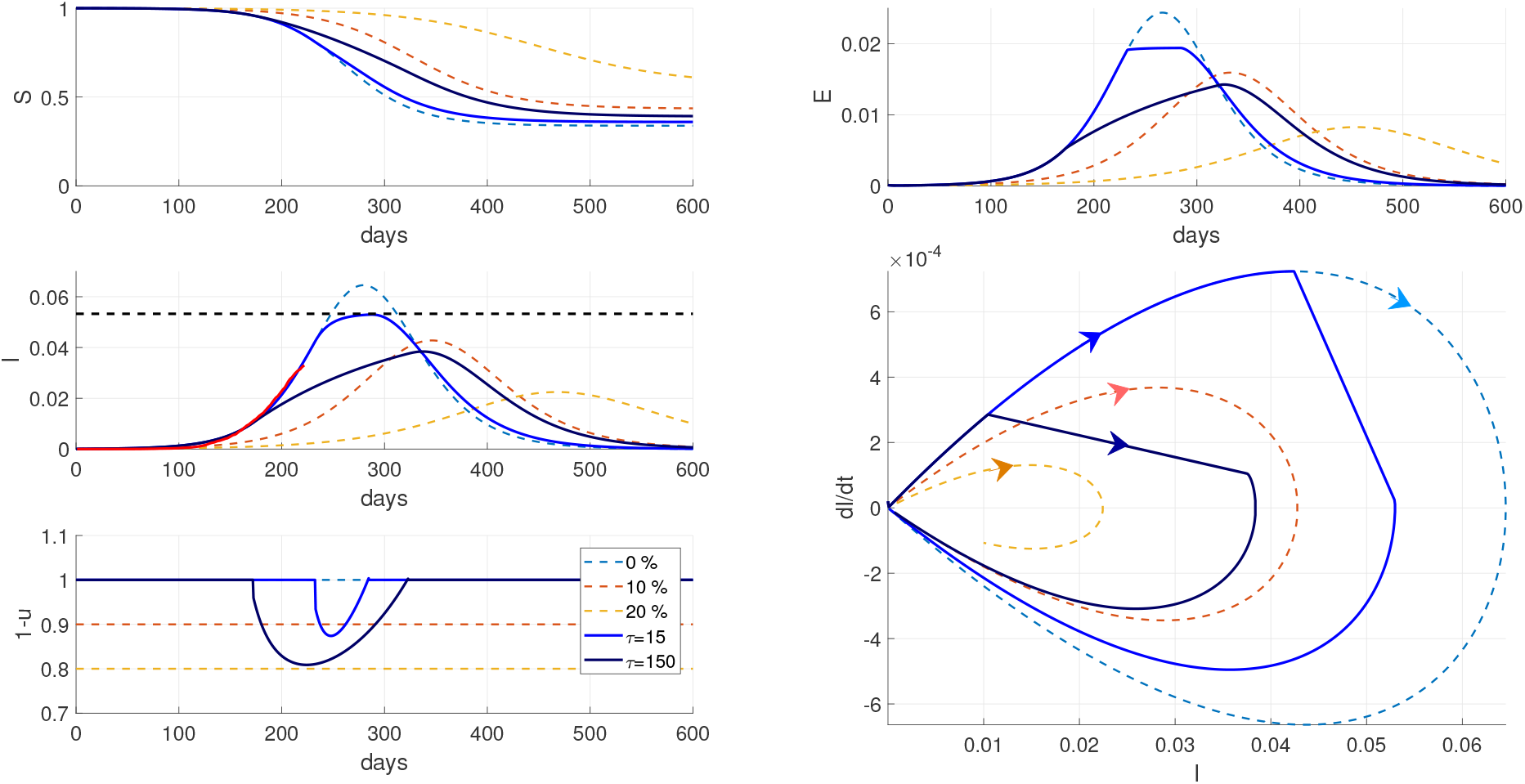
Scenario 2: *β*=0.117 day^−1^. Simulation with SEIR model in a theoretical sce-nario where the switching control action (eq. (4)) is implemented adopting *β*=0.22 day^−1^. From left to right and top to bottom: time evolution of Susceptible (S), Exposed (E), Infectious (I), phase-space plot (I,dI/dt) and restriction actions (1-u). Colored dashed lines represent constant levels of restrictions. Solid light blue and dark blue lines depict the closed-loop evolution for *τ* =15 and *τ* =150 days respectively. Solid red line shows the reported evolution of infectious.

The solid light blue lines in Fig. 4 show the closed-loop evolution for the same limiting conditions than in the first scenario (*I_max_*=1.54e5 and *τ*=15 days). This evolution suggests that an extra level of restriction should be applied to avoid the health system saturation. On the other hand, it can be seen that the proposed strategy would avoid the system saturation without almost extending the epidemic duration. Now, the closed-loop evolution (solid light blue line) differs to a lesser extent with the evolution with u = 0 I (dotted light blue line) due to the restriction and prevention measures already adopted by the government. The solid dark blue lines show the closed-loop system evolution for *τ* = 150 days. Given the possibility of acting before than with *τ* = 15, the maximum infectious limit is never reached, at the cost of some time extension in the epidemic evolution. Table 1 shows the values obtained for the RLI index. As shown, both closed-loop cases (*τ* = 15 and *τ* = 150) requires less additional restriction over the base case, than the 10% one to accomplish for the imposed limit. Besides, the *τ* = 15 case evolution finishes earlier and the τ =150 one achieves a lower level of infectious.

### 4.3 Comparison and discussion

In Fig. 5 a comparison between the two previous scenarios is shown. Using the same plots distribution than before, the free disease evolution (*β*=0.22 day^−1^) is plotted in dashed blue lines. Also, the cases representing a 40% and 47% of constant restriction levels are depicted in dashed orange and yellow, respectively. Again, the solid red line shows the reported real evolution of infectious. As can be seen, the evolution of the disease under the restrictions adopted by the government could be considered equivalent to the evolution for *β*=0.22 day^−1^ with a 47% constant restriction level (dashed yellow lines).

**Figure 5:**
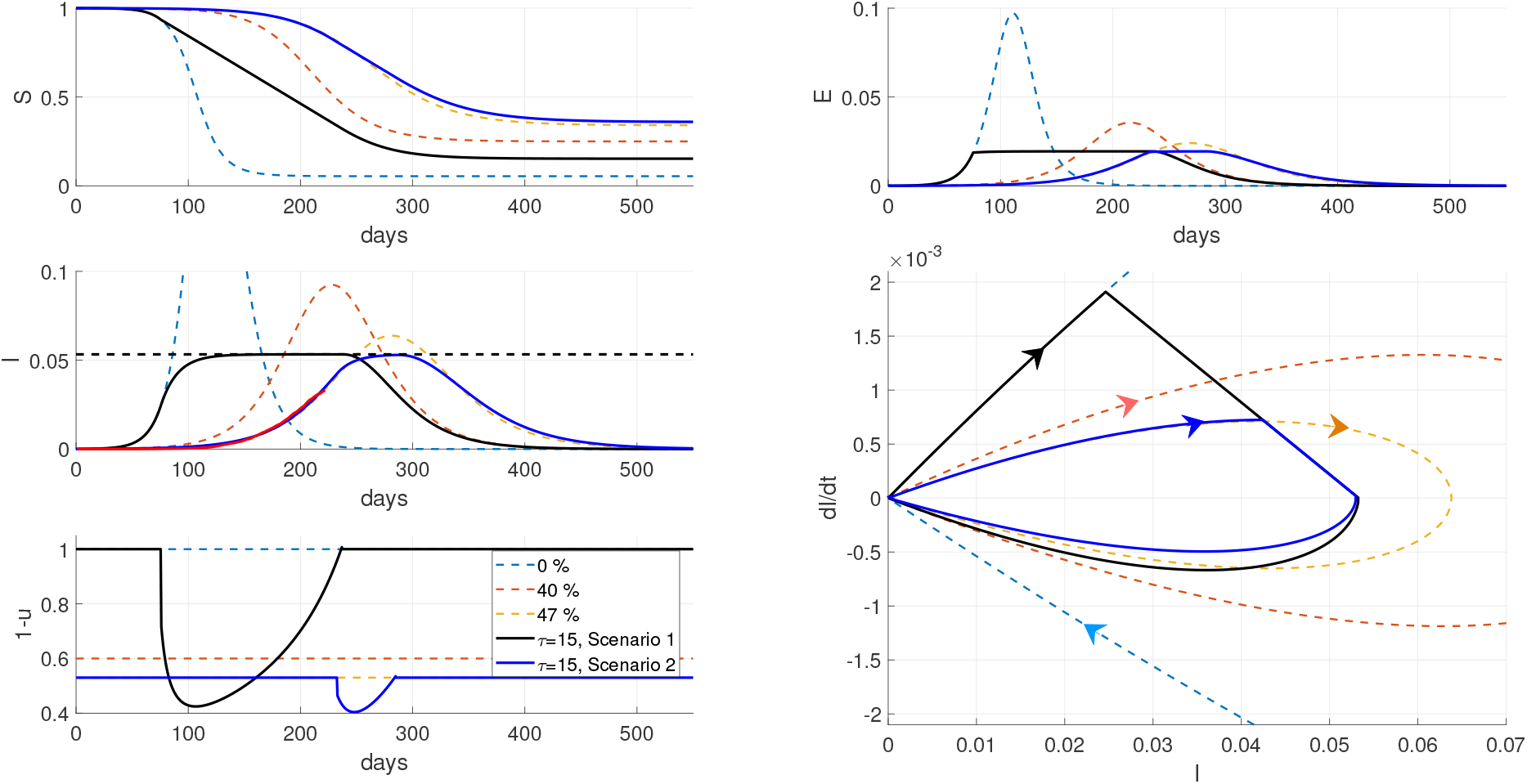
Comparison between scenarios 1 (*β*=0.22 day^−1^) and 2 (*β*=0.117 day^−1^). Simulations with the SEIR model in a theoretical scenario where the switching control action (eq. (4)) is implemented. From left to right and top to bottom: time evolution of Susceptible (S), Exposed (E), Infectious (I), phase-space plot (I,dI/dt) and restriction actions (1-u). Colored dashed lines represent constant levels of restrictions. Solid back lines depict scenario 1 and light blue ones scenario 2 for *τ* = 15 days. Solid red line shows the reported evolution of infectious.

The solid black lines in Fig. 5 show the closed-loop system evolution for the first scenario with *τ* = 15, while the solid lines in light blue depict the closed-loop system evolution for the second scenario using the same τ. As can be seen, the total duration is much lower for the first scenario, that is, applying the SMRC recommendations from the beginning instead of applying more restrictions over the already executed ones by the government. To compare the applied restriction level in both cases, it is necessary to match both scenarios in some way. As mentioned, the second scenario (*β*=0.117 day^−1^ without extra restriction) matches the case of 47 % constant restriction of the first scenario (*β*=0.22 day^−1^). Then, the restrictions which correspond to the second scenario are plotted on top of those corresponding to the 47 % constant restriction, which establishes a “base level” of restriction summarized by *β*=0.117 day^−1^. Thus, the solid light blue line in the lower left box indicates the necessary increase above 47 % restriction, resulting in a much higher level. This can also be seen reflected in Table 1 for the RLI figures. Finally, at the end of the evolution, the first scenario (black case) has a lower number of susceptible than in the second scenario (light blue case) and in the case of 47% constant restriction (yellow) which is beneficial to avoid or reduce a possible second wave of infections.

## 5 Real-world implementation issues

The control action based on a high-frequency switching is not applicable on the population (i.e. continuous opening and closing of economical and social activities is not realizable). The proposed implementation, which makes use of the available measurements (the daily reports of cases), is depicted in Fig. 6. The procedure is summarized in the following steps:

1. Initially, when *I* < *I_max_* the epidemic course is monitored using the available information (new infections, recoveries and deaths) and the function *σ*(*t*) is evaluated. When *σ* = 0 is reached, Step 2 is triggered.
2. The available information is utilized in a model-based simulation (SEIR model + SM algorithm) with a time horizon of T days.
3. From the result of 2) a value is determined to be applied as the NPI. Particularly, the average of the corresponding discontinuous control action, 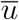, is proposed.
4. The new NPI policy 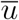 is applied during a fixed period (T days) on the population. Then, proceed to Step 2.

**Figure 6:**
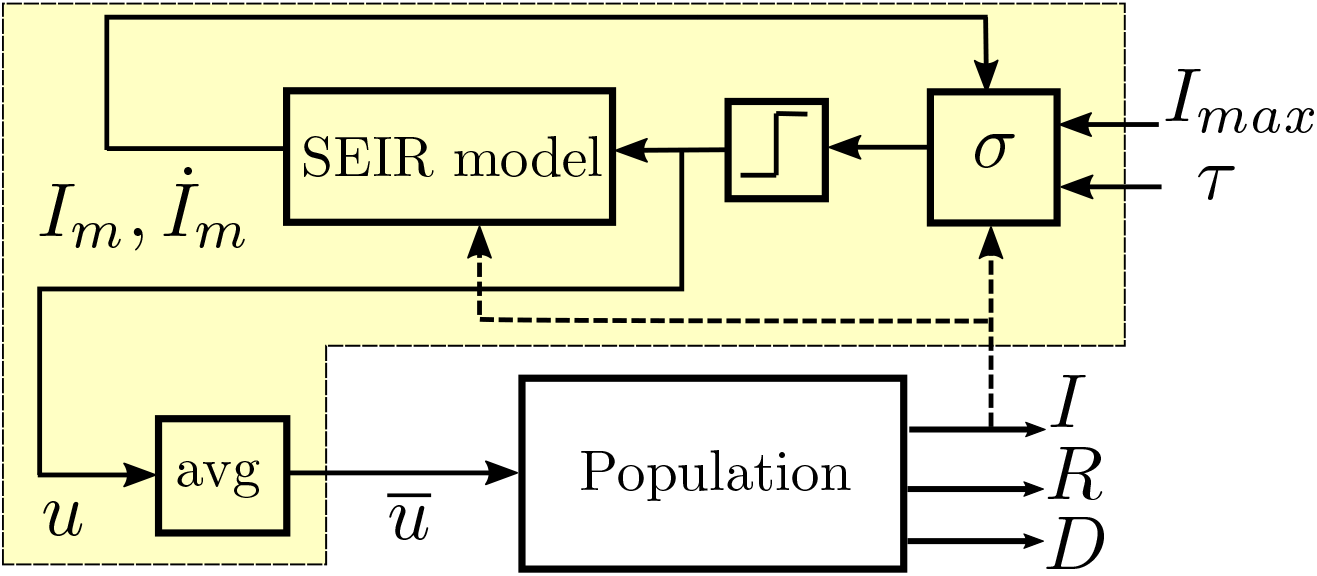
Block diagram of the proposed control scheme. A SEIR model + SMRC conditioning is utilized to periodically update a piece-wise control signal 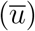.

### Remark

Step 1 acts only on the initial transient and implicitly assumes that the beginning of the intervention can occur in any day of the week. If this is not the case, it can be omitted starting in Step 2.

The information utilized in 2) includes available measurements and it can be extended with updated parameters estimates (e.g. *β, γ*) provided by other algorithms [25]. Also, the value of *I_max_* can be adapted according to the current situation of the health care system. Since the state E is not accessible, the value given in eq. (9) can be taken as the initial condition for E. In Step 3 u can be replaced with a more conservative action (e.g. the maximum value of low-pass filtered version of u). Step 4 describes a basic implementation: once the NPI is issued to the community nothing is done until the period of T days has finished. The proposal can be extended in order to manage specific events such as a sudden soar in the number of infections. Additionally, Step 4 assumes that the control action is effective immediately. It can be adapted taking into account a few days for communicating the new decisions to the population before its effective implementation. Although some important measures (e.g. a lockdown, changes in public transportation rules) require a few days of publishing, there are other actions that can be readily implemented (e.g. number personal transit passes approved per day) [11].

To assess the potential application of the algorithm for the NPI policy design, another significant issue is the translation of u to real-world actions. Certainly any real value in the interval [*U_min_*, *U_max_*] is not realizable. Then, a discretization in a set of finite values should be considered. The mapping between the required values and concrete actions is beyond the scope of this work. Studies in many countries have estimated the effect of different strategies and can serve as a guide for determining the potential effect of each order [2, 27].

### Realistic simulations

A more realistic situation for the first scenario (*β*=0.22 day^−1^) is presented in Fig. 7, where the issues discussed above are taken into account. Black lines correspond to *τ* =15 days and grays lines to *τ* =150 days. Dashed lines represent the theoretical case meanwhile the solid ones do it for the realistic case. Daily sampling of I was considered for calculating *σ*(*t*) during the initial transient. In this case a zero-crossing event is considered for triggering Step 2 and consequently a small delay is introduced. Additionally, in order to incorporate output discretization, only five equally-spaced values in the range [0,1] were allowed. These quantization levels “resemble” the five phases of restrictions adopted by the Argentinian government, phase 1 being the more restrictive one. Each phase contemplates the gradually incorporation of new kinds of activities which could be matched with different constant levels of restrictions. The transition from one phase to another depends on the evolution of the infectious.

**Figure 7:**
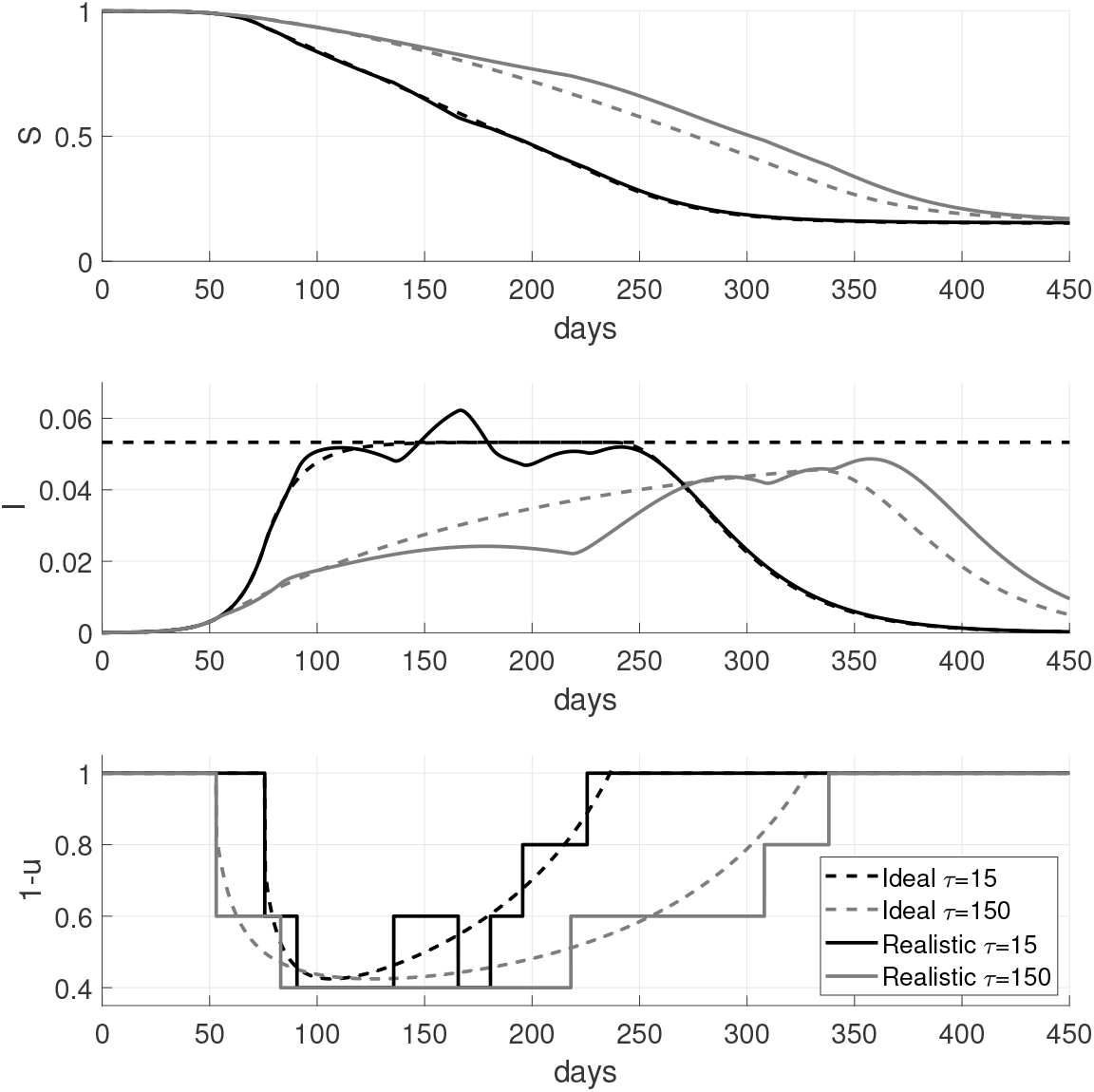
Simulations with the SEIR model by applying a piece-wise control action obtained from a periodic SM algorithm for the first scenario. From top to bottom: time evolution of Susceptible (S), Infectious (I) and applied piece-wise control action (1-u). Black lines are for *τ* =15 meanwhile the gray ones are for *τ* =150. Dashed lines represents the theoretical case and solid lines the discretized ones.

As shown in Fig. 7, the evolution of the realistic scenarios behave very similarly to the ideal ones despite the discretization introduced in the sample time and control action. A little overshoot is shown for the discretized case with *τ* =15 days. This behaviour can be explained as follows. First, the infectious evolution started to decrease (near day 115) and as a consequence the restriction is relaxed (near day 135). But then, with this new fixed restriction level (feature of the discretization) the system evolution surpasses the limit. This behaviour is produced due to the combination of the discretization in a few levels and the use of a smaller *τ*. Further, this value of *τ* coincides with the infectious period 1/*γ*. To avoid the overshoot, the value of τ should be increased. In fact, the*0* 0.5 system evolution for *τ* =150 days (gray lines) puts in evidence the advantage of acting earlier. Due to the control action discretization, the peak level of restriction increments from 58% to 60% which is not a significant increment. These results permit us to show the proposal applicability to the real-world case. Also, it constitutes a useful tool to analyze and compare different hypothetical scenarios to arrive to the best possible solution.

It is important to remark that while in the simulation a given control action is fixed for a period of 15 days, the system evolution can be continuously checked to act in consequence if an improper behaviour is perceived. Within the period of discretization, different types of events may occur in the society, such as protests, which can drastically modify the evolution that was being considered. Also, as a simulation allows one to see future scenarios, different hypothetical situations could be tested to decide which policy to apply at each pandemic stage.

## 6 Conclusions and future research

The COVID-19 pandemic has imposed unprecedented challenges to societies and governments. Pharmaceutical solutions are under development but for the time being, the NPIs are of the most valuable tools to fight the spreading disease. This work assessed the possibility of determining the level of intervention based on sliding mode conditioning ideas and compartmental models.

The proposed algorithm is tuned by three intuitive parameters (*τ*, *I_max_, T*) that provide flexibility to adapt the system response based not only on the number of infectious cases but also on the political and sanitary situation. An improved performance, quantified as lower restriction measures without surpassing the health system capacity, is achievable with the SMRC approach by shaping the infectious time-distribution. The realistic piece-wise discretized control considering a short number of NPI levels performed very close to the theoretical case, showing the proposal applicability.

This SMRC approach could be applied in different cities, where independent levels for *I_max_* can be considered according to the health system capacity and epidemiological situation. Further improvements include periodic update of parameters and coupling with estimators for allowing a time-varying *I_max_*. Also, the application to models that include compartments for quarantine, asymptomatic cases as well as exchange of individuals between regions can be explored.

## Data Availability

Data Availability: see links in References

## Acknowledgments

The authors thanks financial support from MINCYT (BSAS28 COVID-Federal), AN-PCyT (PICT2017-3211), CONICET (PIP0837) and Universidad Nacional de La Plata (UNLP-I253).

